# Protecting healthcare workers during the COVID-19 pandemic: Australian results from the PPE-SAFE survey

**DOI:** 10.1101/2020.08.11.20172148

**Authors:** Mahesh Ramanan, Kevin B Laupland, Alexis Tabah

## Abstract

The COVID-19 pandemic has led to global shortages of personal protective equipment (PPE). Healthcare workers (HCW) have comprised a significant proportion of COVID-19 cases in many countries. The PPE-SAFE survey was conducted to study current practices, availability, shortages, training and confidence in PPE amongst intensive care HCWs around the world. Herein, we describe the results of the Australian respondents to the PPE-SAFE survey. 29% of respondents reported that at least one item of usually available PPE was missing, and 12% reported reuse of single-use items. Only 40% felt that the PPE available to them offered adequate protection. Fit-testing of respirators had never been performed for 47% of respondents, and 49% reported at least one adverse effect from the use of PPE.

---

The COVID-19 pandemic has resulted in over 20 million cases and 700,000 deaths worldwide. Healthcare workers (HCW) comprise upto 11% of cases in some countries(1). The use of personal protective equipment (PPE) may reduce the risk of HCW contracting the infection. Alarming reports of shortages of various types of PPE have emerged along with reports of “homemade” solutions like garbage bags being used as gowns. The PPE-SAFE survey(2) of critical care HCWs from around the world reported on various aspects of PPE usage and HCW safety in reference to caring for COVID-19 patients, including current practices, availability, shortages, training, confidence and HCW adverse effects as reported by HCWs. Herein, we present results from the Australian cohort of PPE-SAFE.

PPE-SAFE was a web-based survey conducted in order to gather HCW-reported data on PPE and PPE-related issues in the context of the COVID-19 pandemic. Participation was voluntary and anonymous. This study was approved and granted a waiver of signed individual informed consent by the Royal Brisbane and Women's Hospital Human Research Ethics Committee (LNR/2020/QRBW/63041).

The survey target population was all HCW of any discipline or training background or level who are directly involved in the management of COVID-19 patients in a critical care setting. A 2-part study-specific survey was designed (see electronic supplement). In the first part, questions surrounding basic demographic, training experience, and institutional work characteristics were elicited. No specific identifying data (i.e. name, date of birth) was requested The second part comprised of a series of questions regarding the usual practices and availability of PPE, along with perceptions of its adequacy in terms of supply and training in the workplace as well as adverse effects of wearing PPE on the HCW. Questions were developed and the survey pre-tested for ease of administration, flow, and content by management committee members and by experienced clinician volunteers. Following iterative revisions, the final survey was developed. An English language version was prepared then translated in the French, Spanish and Italian languages. The survey started with a binary question: if the respondent declared directly caring for COVID-19 patients in the ICU setting the survey was continued and the response categorized as valid. In the opposite case the survey was terminated, and the response categorized as invalid.

The final survey was prepared using the Survey monkey^®^ online platform (SVMK Inc., San Mateo, USA) and posted at https://www.surveymonkey.com/r/PPE-SAFE. The survey was planned to be open for 2 weeks starting March 30. Only the English language version was initially available with the others implemented as of April 7, 2020. Duration of the survey was subsequently extended and we report data collected between March 30 and April 20, 2020.

Subjects were invited to participate through several venues including email invitations using mailing lists of the European Society of Intensive Care Medicine, Australia and New Zealand Intensive Care Society, Australian College of Critical Care Nurses, and the European Society of Clinical Microbiology and Infectious Diseases. In addition, ad hoc emails and advertisements were made via personal networks and social media accounts of management committee members.

Survey results were exported to and analysed using Stata 15.1 (Stata Corp, College Station, USA). Means with standard deviations (SD) and medians with interquartile ranges (IQR)were used to describe normally and non-normally distributed continuous variables, respectively.

The full results of the PPE-SAFE are reported elsewhere(2). 211 (7.8%) of the 2711 respondents were HCWs practicing in Australia, of whom 63% were doctors, 27% were nurses and 10% allied health. Key demographics and usual PPE used are listed in Table 1.

**Table 1:** Demographics of respondents and PPE^1^ used in routine care of COVID-19^2^ patients

|  |  |  |  |
| --- | --- | --- | --- |
| **Demographics** |  | **Mask** |  |
| Age (years) | 44 (35-51)<sup>3</sup> | Surgical Mask | 27, (13%) |
| Female gender | 93, (44%) | N95/FFP2<sup>5</sup> masks | 175, (83%) |
| ICU<sup>4</sup> experience (years) | 13 (7-20)<sup>3</sup> | FFP3<sup>6</sup> mask | 1, (0%) |
| PPE shift duration (hours) | 3 (1-4)<sup>3</sup> | PAPR<sup>7</sup> | 8, (4%) |
| **Position** |  | **Gown** |  |
| Nurse | 56, (27%) | Sleeveless apron | 5, (3%) |
| Physician | 132, (63%) | Full-sleeve waterproof gown | 191, (96%) |
| Allied Health | 23, (11%) | Hazmat suit | 3, (2%) |
| **Usual specialty** |  | **Eye protection** |  |
| Anaesthesia | 3, (1%) | Goggles | 94, (48%) |
| Intensive Care | 200, (95%) | Face shield or visor | 102, (52%) |
| Emergency | 2, (1%) | **Head protection** |  |
| Other | 6, (3%) | Hair cover | 102, (88%) |
| **Hospital type** |  | Balaclava | 12, (10%) |
| Community/urban | 40, (19%) | Impervious hood | 2, (2%) |
| Tertiary | 147, (70%) | **Gloving** |  |
| Private | 7, (3%) | Single | 130, (64%) |
| Remote/regional | 17, (8%) | Double | 72, (36%) |

1 PPE- personal protective equipment
2 COVID-19- coronavirus disease 2019
3 Continuous variables presented as Median (Interquartile range)
4 ICU- intensive care unit
5 FFP2- filtering facepiece 2
6 FFP3- filtering facepiece 3
7 PAPR- powered air purifying respirator

Unavailability of at least one usually available PPE item was reported by 62 (29%) respondents including 14 respondents who reported that N95 respirators were missing. Cleaning and reuse of usually disposable PPE items was reported by 25 (12%) respondents, including N95 respirators (11, 5%) and full-sleeve waterproof gowns (12, 6%). 1 respondent each reported the use of “homemade” cotton masks and gowns. Other individuals reported the use of personal stock of masks, repurposed snorkel masks, use of surgical drapes as hoods and industrial respirators. 40% of respondents felt confident that available PPE available offered them adequate protection.

80% of respondents had received specific PPE training within the last 2 months due to the COVID-19 pandemic. 11% had never had any PPE education and the rest had PPE education at some time prior to COVID-19. Despite this, 59% felt they would benefit from further simulation-based training. 47% had never undergone formal fit-testing of masks, despite recommendations in Australian guidelines(3). 36% underwent fit-testing in the last months due to COVID-19. Only 8% of respondents never used the recommended two-person PPE donning and doffing technique, with one person ensuring that the other dons and doffs correctly without PPE breaches. At least one adverse effect was reported by 49%, including pressure areas (17%), headaches (10%) and thirst (28%).

Concerns about access to adequate PPE is one of the leading causes of HCW anxiety(4). Despite reassurances from Australian authorities, several reports of actual(5) and impending shortages have emerged. Our results from critical care practitioners in Australia show that some of these concerns are not unfounded. Nearly one-third of respondents reported shortages in their institution and 12% reported reuse of single-use items. As PPE-SAFE was a self-reported survey, we were unable to confirm shortages and reuse, and whether reuse was driven by directives or self-initiative. 59% felt they would benefit from further training and almost half had not undergone fit-testing of masks. Only 40% were confident that they were adequately protected. Action is urgently required from healthcare administrators, policymakers, industry and professional bodies to address these concerns.

## Data Availability

Data is available for sharing upon reasonable request to the Chief Investigator of PPE-SAFE, Dr. Alexis Tabah.

## Acknowledgements

We would like to acknowledge the PPE-SAFE investigators and collaborators, and all the healthcare workers who took the time to fill out our survey in the midst of a severe crisis.

